# Estimated genetics prevalence of early-onset Parkinson’s disease caused by *PRKN* mutations

**DOI:** 10.1101/2024.01.22.24301610

**Authors:** Dorothée Diogo, Emily H.M. Wong, Christopher DeBoever, Wei Qu, Jonghun Lee, Stephen Crawford, Takeshi Hioki, Jaya Padmanabhan, Daria Prilutsky, Gabriele Proetzel

## Abstract

**Background:** Estimating the prevalence of rare diseases is challenging due to very limited natural history studies, lack of studies in diverse populations, and frequent under or misdiagnosis. We leveraged human genetics to estimate the genetic prevalence (eGP) of familial Parkinson’s disease (PD) caused by biallelic pathogenic variants in the *Parkin* (*PRKN*) gene (PRKN-PD).

**Methods:** We curated the reported *PRKN*-PD pathogenic variants and obtained the heterozygous carrier frequencies of these variants from gnomAD and the Japanese Multi-omics reference panel (jMorp). We used the carrier frequencies to estimate the eGP of *PRKN*-PD in eight genetic ancestries.

**Results:** Non-Japanese East Asians presented the highest eGP of *PRKN*-PD (24 per 100,000 individuals, 95% CI=4-165 per 100,000 individuals), followed by Non-Finnish Europeans (22 in 100,000 individuals, 95% CI = 11-64 per 100,000 individuals). Based on the proportions of races and ethnicities, we estimated the eGP in the USA and the world-wide eGP to be 18 per 100,000 individuals (95% CI=7-68 per 100,000 individuals). and 13 per 100,000 individuals (95% CI=3-70 per 100,000 individuals), respectively. These estimates were significantly reduced when excluding structural variants (world-wide eGP=2 per 100,000 individuals, 95% CI=1-5 per 100,000 individuals).

**Conclusions:** This is the first study estimating the *PRKN*-PD genetic prevalence. Our results suggest that the prevalence of the disease may be higher than previously reported, highlighting potential underdiagnosis. We also demonstrate the importance of carefully considering the known genetic epidemiology of each disease, and its limitations, when using the approach applied in this study to estimate the disease genetic prevalence.

## Introduction

As of 2022, about 7000 rare diseases have been identified^1^. Collectively, rare diseases affect around 25– 30 million people in the United States (US) and more than 300 million people worldwide. Only 5% of rare diseases have FDA-approved drug treatments, highlighting the high unmet need remaining for patients living with these diseases^2^.

Robust epidemiology data is critically important to better assess the burden of a rare disease on patients and families^3^. In drug development, this information is crucial to help guide clinical development strategies. Traditionally, one can measure how common a disease is in a population by tallying the number of diagnosed patients in the population itself. However, many rare diseases are under-diagnosed, leading to unreliable disease prevalence estimates. For rare autosomal recessive (AR) diseases, where pathogenic variants in both alleles (maternal and paternal) are required to cause the disease, the frequency of known pathogenic variants in large reference population datasets serve as a valuable metric to estimate how common a disease may be. This approach estimates the genetic prevalence, i.e., the proportion of people with specific causal pathogenic genotypes in the population. Assuming full disease penetrance of the variants, the genetic prevalence refers to the lifetime risk – the probability of an individual (from birth) to eventually developing the disease. Several published studies have used this approach to predict the genetic prevalence of rare recessive diseases, including, for example, Pompe disease, Wilson disease, inherited retinal diseases, ENPP1 deficiency and congenital thrombotic thrombocytopenic purpura^4–8^.

Familial Parkinson’s disease (PD) is an autosomal recessive (AR) condition caused by pathogenic variants in the *Parkin* (*PRKN*) gene^9^. *PRKN* is located on chromosome 6q25.2-27, contains 12 exons and is one of the largest human genes. It encodes an E3 ubiquitin ligase critical in targeting substrate proteins for proteasomal, and is one of the two kinases that has a mitochondrial targeting for regulating mitochondrial quality control and promoting the selective autophagy of depolarized mitochondria (mitophagy)^10^. Biallelic loss-of function variants in *PRKN* cause autosomal recessive familial Parkinson’s Disease^9^. Clinically, *PRK N*-PD manifests with early disease onset (median age at onset being 31 years old), slowly progressive and classical motoric symptoms, including bradykinesia, resting tremor, rigidity, and clinically and pathologically distinct from idiopathic PD^11^. Neuropathologically, it is differentiated by general absence of abnormal α-synuclein deposition, incidental Lewy body pathology and mild neuronal loss confined to the substantia nigra and locus coeruleus^12^.

*PRKN*-PD has been suggested to represent 0.5-1% of individuals with PD across all age-at-onset (AAO) groups based on the proportions of *PRKN* pathogenic variants carriers observed in published PD studies^13^. Together with the estimated total number of PD patients in the world^14^, the number of *PRKN*-PD patients is estimated at 35,000-70,000 worldwide^13^. *PRKN* pathogenic variants are more prevalent among adolescent PD patients (AAO<20 years old, 77%) and adult patients with AAO<=50 years old (2.6-2.8%)^15–17^. However, prevalence estimates from these studies might be affected by ascertainment bias, including different proportions of familial and idiopathic patients recruited, and small sample sizes. In addition, ancestral background of patients in these studies, which could associate with differential variant frequencies, is often not reported or accounted for. Therefore, the prevalence estimated by these studies might not be reliable.

In this study, we estimated the genetic prevalence (eGP) of PRKN-PD in diverse ancestries using heterozygous carrier frequencies of reported pathogenic *PRKN* variants, including small variants (single nucleotide variants and frameshift-causing small indels, referred below as SmVs) and structural variants (SVs), We also estimated eGP of PRKN-PD in the USA, based on the estimates obtained in the different ancestries and the proportions of sub-populations reported in the US census.

## Methods

### Compilation of reported pathogenic **PRKN** variants from different data sources

We queried pathogenic variants information from three public databases – MDSGene, ClinVar and OMIM (**Figure 1**)^18–20^.

**Figure 1:**
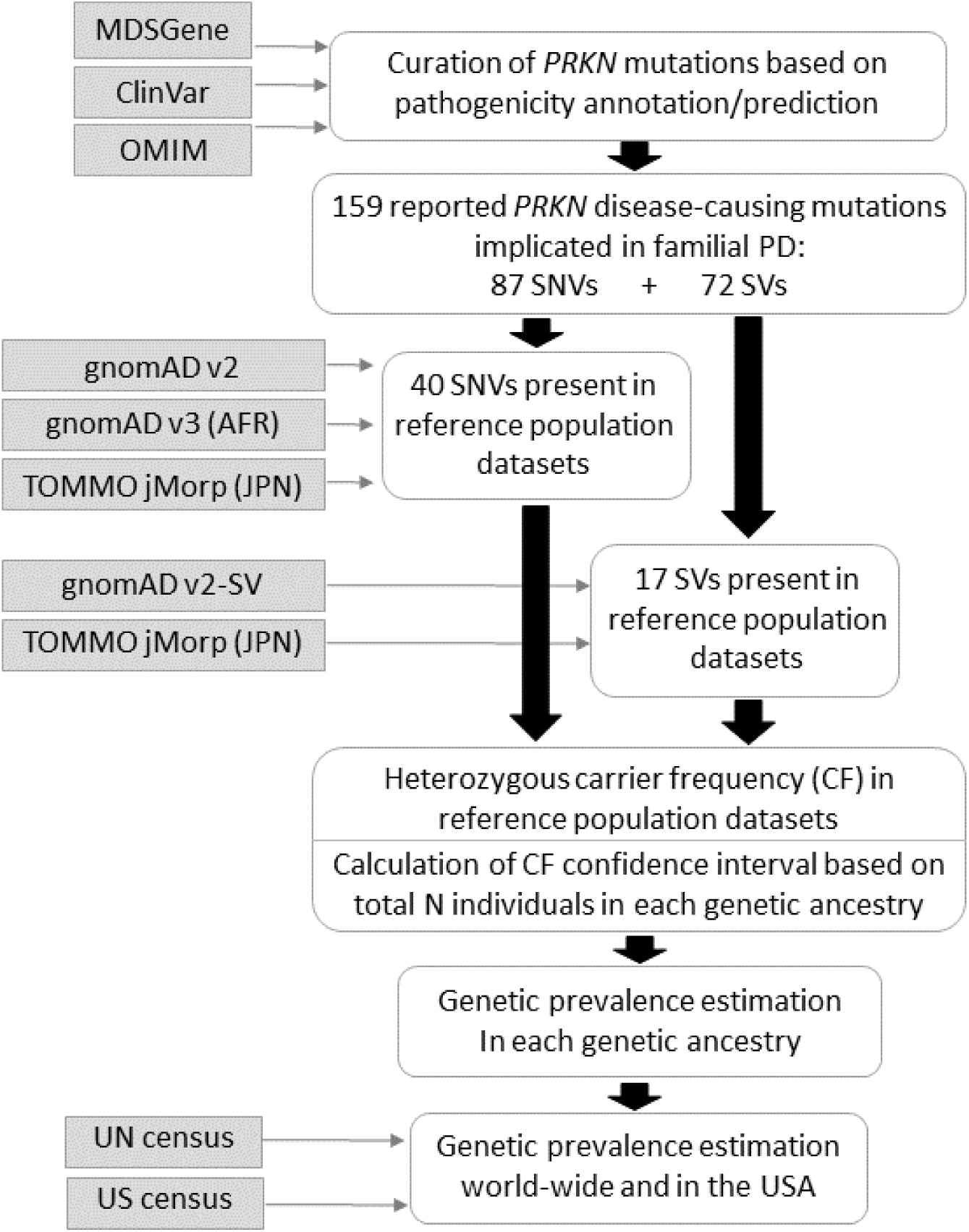
Study design overview. We identified 159 known pathogenic variants in *PRKN* implicated in autosomal recessive *PRKN*-Parkinson disease (*PRKN*-PD), out of which 57 were observed in the reference population databases gnomAD or jMorp and were included in our genetic prevalence estimations. SmVs, single nucleotide variants and frameshift-causing small indels ; SVs, structural variants.

MDSGene performed a systematic literature screening and curated phenotypic and genotypic data of *PRKN*-PD patients from 174 studies^11,18^. We obtained the curated genotypes and ages at disease onset (AAO) of *PRKN*-PD patients from the MDSGene online database (www.mdsgene.org, last accessed on 3 August 2022).

We obtained *PRKN* variants reported in ClinVar using the website https://www.ncbi.nlm.nih.gov/clinvar/ (last accessed on 9 December 2021). We only considered pathogenic or likely pathogenic variants implicated in PD that were rated with at least two gold stars (including review status “criteria provided, multiple submitters, no conflicts” (2 gold stars), “reviewed by expert panel” (3 gold stars) and “practice guideline” (4 gold stars)).

We extracted *PRKN* small variants that were reported in OMIM (https://omim.org/) to be causing autosomal recessive PD (last accessed on 13 December 2021).

Finally, we added the variant c.535-3A>G which has recently been reported in three Japanese families ^21^.

For each reported pathogenic mutation, we assessed the evidence from the three mutation databases, and we excluded those with conflicting evidence – reported as pathogenic in one source while benign in another. We further assessed the pathogenicity of these mutations based on ACMG guidelines^22^. We inferred the GRCh38 genomic coordinates of these mutations, when not available, based on the HGVS nomenclature or the GRCh37 genomic coordinates.

### Reported **PRKN** pathogenic variants observed in human reference databases

To obtain allele frequencies (AF) of the *PRKN* pathogenic variants in the general population, we interrogated five data sources (**Figure 1**).

We interrogated gnomAD v2.1.1 to obtain allele frequencies of SmVs in most ancestries, except African/African American and Japanese ancestries. For allele frequencies of SmVs in African/African Americans, we interrogated gnomAD v3. For allele frequencies of SmVs in Japanese, we interrogated WGS data generated by the Tohoku Medical Megabank Organization (ToMMO) via the Japanese Multi-Omics Reference Panel (jMorp) website^23^. Both the gnomAD v3 African/African American cohort and the Japanese cohort in JMorp include a substantially larger number of individuals compared to gnomAD v2, thus providing more accurate allele frequencies in those populations.

GnomAD v2 SmVs: We obtained the gnomAD v2.1.1 liftover SmV data from the gnomAD website (https://gnomad.broadinstitute.org/downloads, last assessed on 20 July 2023). This data was generated from 141,456 unrelated individuals from different ancestral groups (125,748 whole exomes and 15,708 whole genomes), lifted over from the GRCh37 to the GRCh38 human reference sequence.

GnomAD v3 SmVs: We obtained the gnomAD v3.1.2 SmV allele information from whole genome sequences of 20,744 individuals of African genetic ancestry. All the genome sequences were mapped to GRCh38.

TOMMO SmVs: We accessed allele frequencies of SmVs in Japanese ancestry from TOMMO (https://jmorp.megabank.tohoku.ac.jp/). These summary statistics were generated from whole genome sequencing data of 54,000 Japanese individuals.

In order to incorporate structural variant population data, we leveraged two data sources. We first obtained the SV data recently made available from 10,847 genomes in gnomAD v2.1^24^. We also leveraged SVs information from WGS of 8,300 Japanese available on the TOMMO jMorp^25^. We linked the SVs reported in gnomAD v2.1 and TOMMO-8.3KJPN-SV to variants found in PRKN-PD patients based on the genomic region overlap. We assumed that the SVs were the same if they were of the same variant type (deletion or duplication) and had the same consequences on the cDNA (e.g. complete deletion of exon 3).

**Supplementary Table 1** shows the number of sequenced individuals in each database.

### Calculation of heterozygous pathogenic variants carrier frequencies

We inferred the frequencies of heterozygous pathogenic variants carriers (individuals not affected by early onset PD), also known as “carrier frequency”, in different ancestral populations, using the human reference databases mentioned above.

For each reported pathogenic *PRKN* variant observed in gnomAD v2, gnomAD v3 or TOMMO, we calculated the carrier frequency in each population using the following formula:

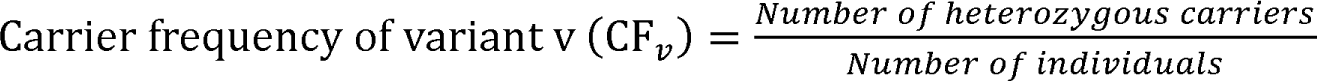

For each variant and each ancestry population, we calculated the 95% confidence interval (CI) of the CF using the Wilson score.

### Estimation of genetic prevalence in various ancestries

To estimate the genetic prevalence of *PRKN*-PD in each ancestral population, we used the following approach as described elsewhere^8^.

Assuming all biallelic variant combinations were possible, pathogenic and fully penetrant, we considered all possible combinations of two known pathogenic variants in the calculation of *PRKN*-PD genetic prevalence. Considering each possible pair of variants i and j (where i and j can be either the same variant, i.e. homozygote genotype, or different variants, i.e. compound heterozygote genotypes), we estimated the genetic prevalence in each ancestry using the formula:

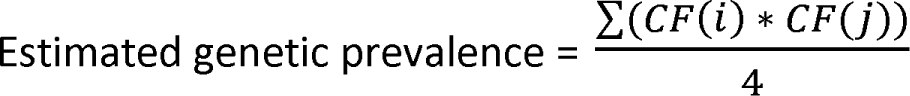

This estimated genetic prevalence of *PRKN*-PD is equivalent to the probability of heterozygous carriers having offspring with biallelic pathogenic variants.

We used the CI lower bounds and upper bounds of the variants’ CF to calculate the genetic prevalence CI in each ancestry population.

### Inference of *PRKN*-PD genetic prevalence in the USA and world-wide

We inferred the *PRKN*-PD genetic prevalence in the USA based on 1) the estimated PRKN-PD genetic prevalence in the different ancestries, and 2) the major population proportions reported in the US census as well as the proportions of European and Asian sub-populations reported in the community surveys.

The proportions of major populations in the USA were obtained from the 2021 US Census data (https://www.census.gov/quickfacts/fact/table/US/PST045221, last accessed on 20 July 2022). We also obtained the proportion of self-reported Europeans and Asians from the American Community Survey website (https://www.census.gov/programs-surveys/acs/data.html).

We matched these self-reported races and ethnicities or countries of origin to the genetic ancestries defined by the human reference population databases and inferred the *PRKN*-PD genetic prevalence in the USA. We applied the same approach, using the 2022 Revision of World Population Prospects published by the United Nations (https://population.un.org/wpp/), to estimate the worldwide genetic prevalence of PRKN-PD.

## Results

### Reported pathogenic *PRKN*-PD variants

We first compiled and curated the reported human pathogenic *PRKN*-PD variants from three databases (MDSGene, ClinVar, OMIM), and investigated their allele frequency in five large diverse population datasets (gnomAD v2, gnomAD v2-SV, gnomAD v3, ToMMO-54K and ToMMO-8.3K-SV) (**Figure 1**).

A total of 159 distinct disease-causing genetic variants were identified in the *PRKN* gene, after consolidating structural variants (SV) with same consequences on coding sequence (**Supplementary Table 2**). Among these, 72 (45.3%) were SVs (large deletions, large duplications, large indels, other large rearrangements), 48 (30.2%) were predicted loss-of-function (pLoF) single nucleotide variants (SmVs: nonsense, frameshift, or splice variants), 33 (20.8%) were missense variants and 6 were other types of SmVs.

Out of the 159 disease-causing *PRKN* variants, 57 were present in the population datasets. This includes 17 SVs (12 large deletions and 5 large duplications) present in gnomAD v2.1 or ToMMO, and 40 SmVs present in gnomAD v2, gnomAD v3 or ToMMO.

We interrogated MDSGene^18^, which provides patient-level genotype and phenotype data, to investigate the frequency of individual variants among *PRKN*-PD patients, identify the variants most frequently reported in patients and ensure that those variants were present in the population datasets to contribute to the genetic prevalence estimations.

As previously described^11^, the most common variant in *PRKN*-PD patients in MDSGene was c.(171+1_172-1)_(412+1_413-1)del, resulting in a complete deletion of exon 3. This SV was observed in 99 patients out of 701 (14%) index *PRKN*-PD patients. This variant was more frequent in patients of reported Asian ethnicity: 58.7% *PRKN*-PD index patients with this variant were of Asian ethnicity, in contrast to 32.7% Asians reported among *PRKN*-PD index patients not carrying this SV (P=7×10^-4^). Six additional SVs leading to complete deletion of exon 3 were reported in seven Caucasian homozygous or compound heterozygous *PRKN*-PD index patients, bringing the total number of index patients with an exon 3 deletion to 106 (15%). In gnomAD v2.1, nine distinct SVs causing complete deletion of exon 3 were reported. The most frequent of those nine SVs (DEL_6_76253) had an allele frequency (AF) ranging between 0.01% in European and African/African American ancestries, and 0.08% in East Asians. When combining all nine SVs reported in gnomAD v2.1, the allele frequency of exon 3 deletions across different populations was 0.07%, ranging between 0.04% in African/African Americans, 0.08% in Europeans, 0.12% in East Asians and 0.16% in Latino/admixed American ancestries. Two SVs corresponding to a complete deletion of *PRKN* exon 3 were reported in the ToMMO 8.3K WGS data with an aggregated allele frequency of 0.04%.

The second most common pathogenic variant was the frameshift variant c.155delA (p.Asn52Metfs*29), reported in 78 (11%) *PRKN*-PD index patients. Most patients (92.1%) carrying this variant were Caucasians (in contrast to 40.9% Caucasians reported in the overall PRKN-PD index patients cohort, P=3.6×10^-12^). In the general population (based on gnomAD v2.1.1, gnomAD v3, and JMorp), this frameshift variant was most frequent among Latino or admixed Americans (AF=0.11%) followed by non-Finnish Europeans (AF=0.024%) and African/African Americans (AF=0.012%). This variant was absent in Asian reference population cohorts (AF=0, in gnomAD and ToMMO).

### Estimated genetic prevalence of *PRKN*-PD in the general population

We next leveraged the allele frequencies of the 57 *PRKN* known disease-causing SmVs and SVs present in the population datasets to estimate the genetic prevalence (eGP) of *PRKN*-PD in four major genetically-defined populations in gnomAD, and in Japanese using the ToMMO data (**Figure 2** and **Supplementary Table 3**). We also calculated the genetic prevalence in six non-Finnish European (NFE) subpopulations, in two East Asian genetically-defined subpopulations, in Ashkenazi Jews, and in South Asians represented in gnomAD v2 (**Supplementary Table 3**). However, we note that some (sub)populations eGP are based only on SmVs and do not include SVs information due to absence of SV reference data in those populations.

**Figure 2:**
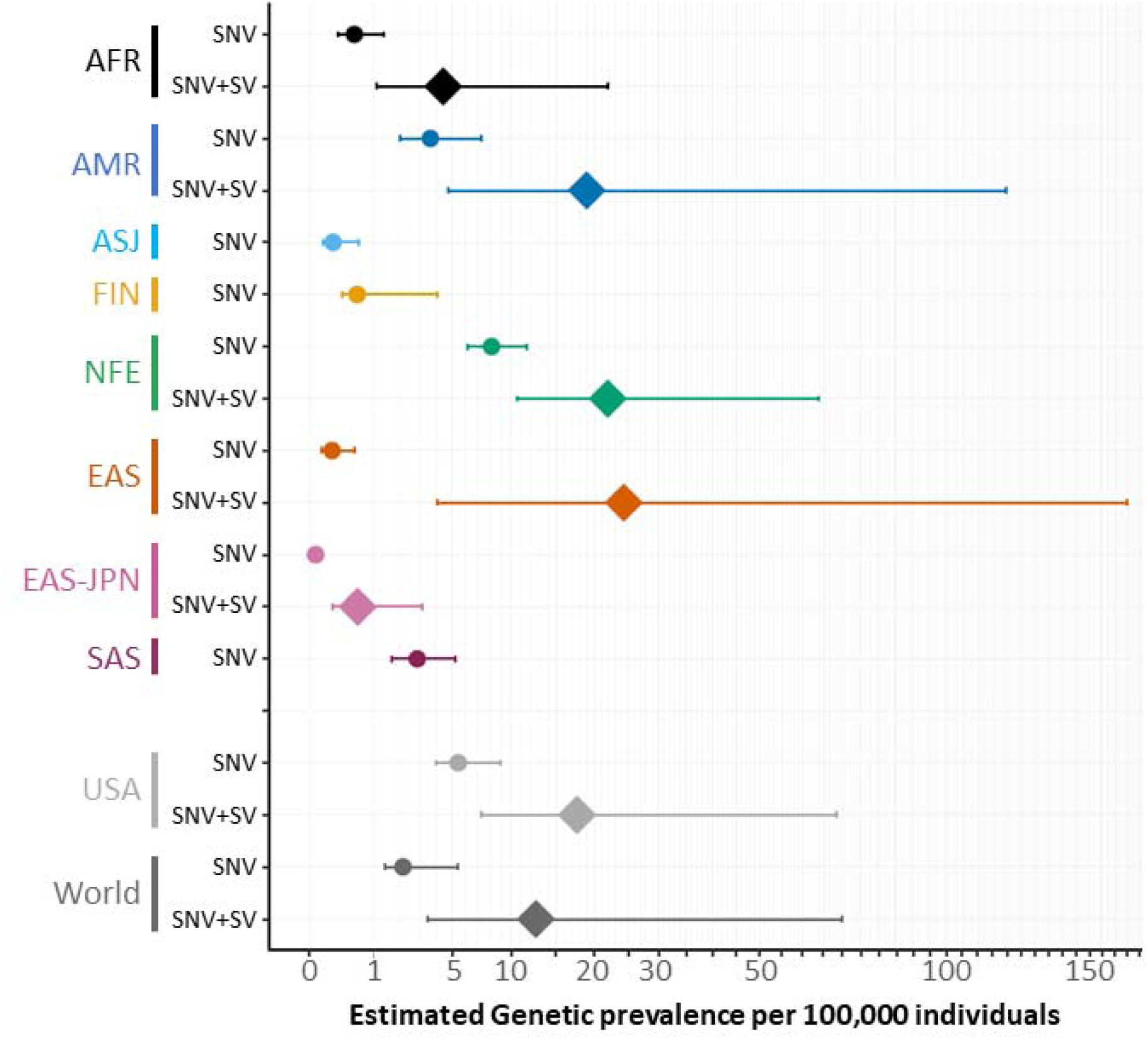
Genetic prevalence estimates in various genetic ancestries, in the USA and world-wide. The x-axis indicates the estimated genetic prevalence eGP (and 95% confidence interval, Wilson score) per 100,000 individuals (Log2 scale). For each major genetic ancestry, two sets of eGP are shown: eGP based on SmVs (SmV), eGP based on SmVs and SVs (SmV+SV). World-wide eGP and eGP in the USA (SmV and SmV+SV) were calculated based on the estimates in each genetic ancestry and the proportion of each ancestry in the UN and US census. AFR, African/African American ; AMR, Latino/Mixed American ; ASJ, Ashkenazi Jewish ; EAS, East Asian ; EAS-JPN: Japanese ; FIN, Finnish ; NFE, Non-Finnish European ; SAS, South Asian. SmV, single nucleotide variants and frameshift-causing small indels; SV, structural variant.

As shown on **Figure 2** , East Asians had the highest eGP (24 in 100,000 individuals, 95% CI=4-165 per 100,000 individuals), largely driven by SVs, particularly deletions of exon 2, exon 3 and exons 2-3 (**Figure 3**).

**Figure 3:**
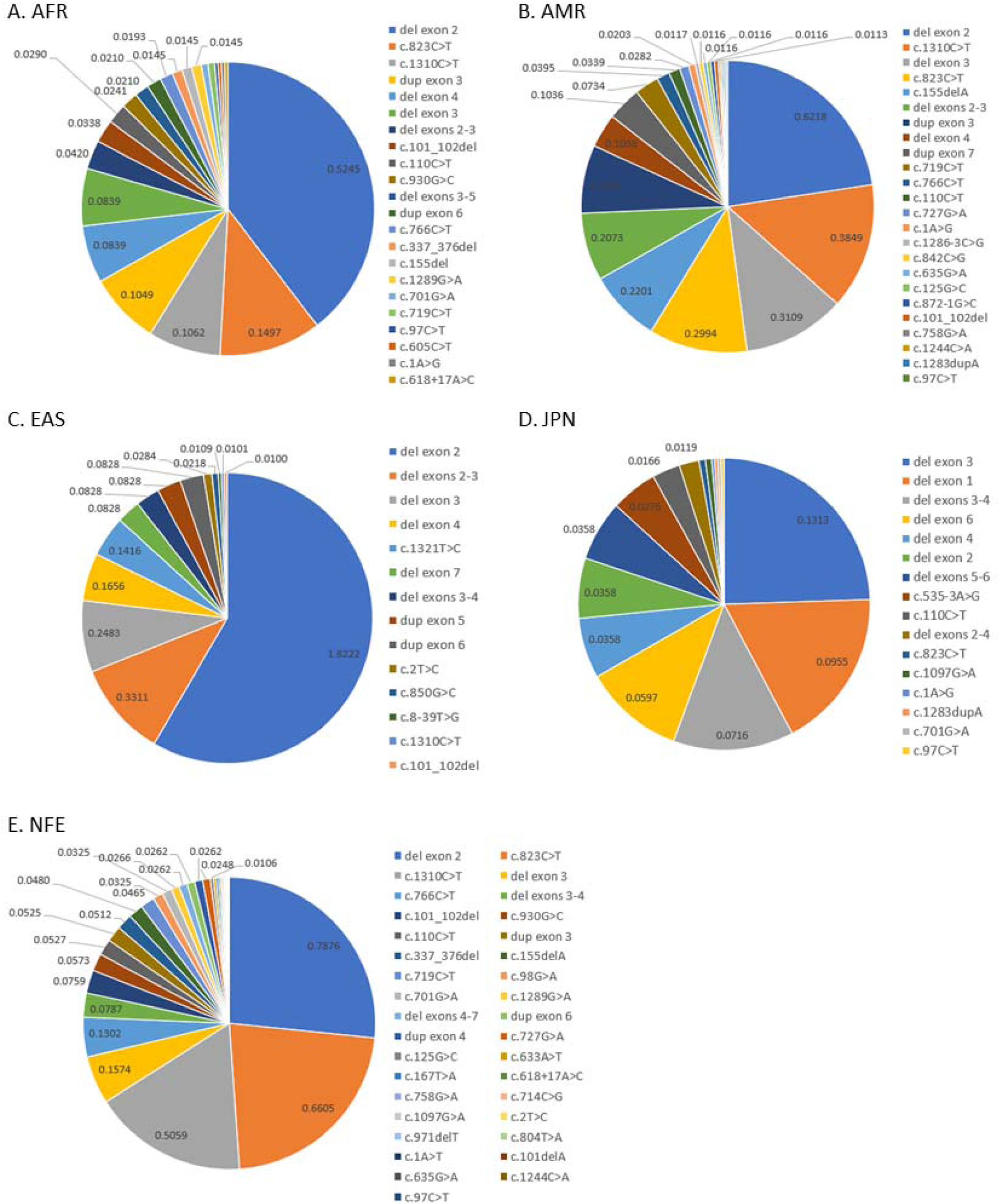
Heterozygous carrier frequency of pathogenic *PRKN* single nucleotide variants (SmVs) and structural variants (SVs) in African/African Americans (A), Latino/admixed Americans (B), Non-Japanese East Asians (C), Japanese (D) and Non-Finnish Europeans (E).

The second highest eGP was observed in NFE (22 in 100,000 individuals, 95% CI=11-64 per 100,000 individuals). We note that NFE had the highest lower bound eGP (11 in 100,000 NFE individuals vs 4 in 100,000 East Asians). The three most frequent variants contributing to eGP in NFE included deletions of exon2, c.823C>T (p.Arg275Trp) and c.1310C>T (p.Pro437Leu) (**Figure 3**). The two SmVs were also among the top five most frequent variants contributing to eGP in all other non-Asian populations (**Figure 3**). c.823C>T and c.1310C>T were reported in 68 (9.7%) and 13 (1.9%) index patients in MDSGene, respectively, and predominantly in Caucasians (85.7% and 100% of index patients with those pathogenic variants and ethnicity were Caucasians, respectively).

The lowest eGP was observed in Japanese, with eGP=6 per 1 million individuals (95% CI = 1 per 1 million-3 per 100,000 individuals) and was largely driven by SVs as well as two SmVs (c.535-3A>G and c.110C>T/ p.Pro37Leu). (**Figure 2** , **Figure 3** and **Supplementary Table 3**). Because our Japanese estimates are derived from a large cohort of individuals (thus enhancing the accuracy of the carrier frequencies), and because of the small number of individuals of Japanese ancestry within the East Asian cohort in gnomAD v2 (N=76 Japanese out 9,977 East Asians in the gnomADv2 SmV dataset; unknown number of Japanese out of 1,208 East Asians in gnomADv2 SV dataset), the high eGP in East Asians is likely driven by high eGP in non-Japanese East Asians.

We next determined the genetic prevalence of *PRKN*-PD in the USA by extrapolating from the reported proportions of races and ethnicities in the US Census and American Community Survey, coupled with the estimated genetic prevalence of the corresponding genetically-defined populations described above. Based on this analysis, we estimated the genetic prevalence of *PRKN*-PD in the USA to be 18 per 100,000 individuals (95% CI: 7-68 per 100,000 individuals) (**Figure 2** and **Supplementary Table 3**).

Using the same approach and the proportion of races and ethnicities in the 2022 Revision of World Population Prospects published by the United Nations, we estimated the world-wide genetic prevalence of *PRKN*-PD to be 13 per 100,000 individuals (95% CI: 3-70 per 100,000 individuals) (**Figure 2** and **Supplementary Table 3**).

## Discussion

In the literature, at least 1,530 symptomatic *PRKN-*PD patients have been reported to date^26^. Based on previously published studies, Wasner et al suggested that *PRKN*-PD represents around 0.5–1% of PD patients across all AAO groups^13^. Based on the data from the Global Burden of Disease Study, Dorsey et al estimated that around 6.2 million individuals worldwide have PD, representing 0.082% of the total population (7.6 billion)^14^. Assuming *PRKN* biallelic pathogenic variants contribute to 0.5–1% of PD patients across all age groups, the prevalence of *PRKN*-PD would be 4-8 in 1 million individuals worldwide.

*PRKN*-PD, like many other rare diseases, is better understood in specific human populations where the majority of studies have been conducted. In MDSGene, the majority of *PRKN*-PD patients are of Asian (39%; primarily from Iran, China or Japan) or Caucasian (31%) ancestry^11^. In order to better understand the genetic epidemiology of *PRKN*-PD in diverse populations, we have investigated the *PRKN*-PD genetic prevalence eGP within eight genetically-defined ancestries and eight genetically-defined subpopulations by leveraging large-scale genetic datasets from five human reference population databases.

When estimating the genetic prevalence of a disease, it is imperative to first meticulously curate the reported disease-causing variants from confirmed patients, carefully investigate the frequency of the known pathogenic variants and variants types in reported patients, and take into account the ancestries represented within the cohort of reported patients, in order to inform the eGP analysis design and interpretation of results. Previous GP studies in rare diseases have largely focused on diseases predominantly caused by protein-coding SmVs and relied on gnomAD as the only source of SmV carrier frequencies. Our study design differs from previous GP studies on two fronts. First, in order to enhance the accuracy of the SmV carrier frequencies, we harnessed data from three reference databases (gnomAD v2, gnomAD v3 and jMorp), and selected for each variant and each ancestry, the carrier frequency from the reference database with the largest sample size. Notably, the inclusion of jMorp enabled the utilization of carrier frequency data from 54,000 Japanese, an ancestry that is significantly underrepresented in gnomAD. Secondly, as described by MDSGene and other publications, SVs contribute significantly to the genetic cause of familial *PRKN*-PD ^11,21^. To address this observation, we probed two reference datasets – gnomAD v2-SV and ToMMO 8.3K SV - that provide carrier frequency of SVs, and highlight the role of SVs in the genetic prevalence of *PRKN*-PD.

Three key findings can be derived from our results. First, our results suggest that the prevalence of *PRKN*-PD may be substantially higher than previously thought, with an eGP of 13 per 100,000 individuals worldwide (95% CI: 3-70 per 100,000 individuals) (**Figure 2** and **Supplementary Table 3**). Second, our results indicate that *PRKN*-PD eGP varies significantly between ancestries ranging from only 6 in 1 million Japanese (95% CI = 1 per 1 million-3 per 100,000 individuals) to 22 in 100,000 NFE (95% CI=11-64 per 100,000 individuals) and 24 in 100,000 non-Japanese East Asians (95% CI=4-165 per 100,000 individuals). Third, our analysis supports that the variants types contributing to the disease etiology varies greatly between ancestries, with the highest frequency of disease-causing SVs in non-Japanese East Asians and highest frequency of disease-causing SmVs in NFE (**Figure2** and **Figure 3**). Interestingly, while the *PRKN*-PD literature includes a significant proportion of patients from Japan^11,21^, our estimates suggest that the genetic prevalence of the disease is low in the Japanese population. The majority of the Japanese index patients reported in MDSGene carry large deletions within *PRKN* (80/86 alleles, 93%), particularly deletions of exon 3, exon 4 and exons 3-4 (50/86 alleles, 58%). Those three deletions were present in the TOMMO 8.3KJPN-SV dataset and contributed to our genetic prevalence estimate with an aggregate heterozygous carrier frequency of 0.18% (**Figure 3**). A recent study reporting pathogenic variants in a large Japanese familial PD cohort reported the pathogenic SmV c.535-3A>G in multiple families^21^. This splice variant was the most frequent pathogenic SmV in the TOMMO 54K dataset and contributed to our genetic prevalence estimate with a carrier frequency of 0.28% (**Figure 3**). Our findings suggest that the high proportion of Japanese patients described in the literature may be due to ascertainment bias and may not reflect the percentage of Japanese patients in the global patient population. Even larger, high-quality, reference SV frequency data in Japanese will be required to validate this finding.

It is important to make the distinction between the genetic prevalence, which reflects the proportion of individuals that carry two pathogenic variants, and the disease prevalence, which reflects the proportion of symptomatic patients. If pathogenic variants are not fully penetrant (i.e. not all individuals carrying a biallelic genotype with the variant will develop symptoms), the genetic prevalence and the disease prevalence may be inconsistent. In the case of *PRKN*-PD, no robust information about variant penetrance is known. A recent study in UK Biobank reported that, while carriers of two *PRKN* pathogenic variants were more likely to have a Parkinson’s disease diagnosis, 22 (91.7%) biallelic carriers had no symptoms^27^. This observation can represent incomplete ascertainment of disease status, incomplete information due to difference between current age of the individuals and age of disease onset, and/or can reflect incomplete penetrance of (some of) the variants. Noteworthily, deletions of exons 2, 3 and 4 have been reported with a frequency higher than expected in the general population for pathogenic variants. In the gnomAD v2 dataset (N=10,847 samples), SVs leading to complete deletions of exon 2, exon 3 or exon 4 (which were reported in 57, 99 and 67 homozygous and compound heterozygous index patients in MDSGene) have an aggregate carrier frequency of 0.8%, 0.15% and 0.06% respectively in gnomAD v2 (**Figure 3**). More work will be required to better assess SVs in *PRKN* (and gene-level SV status per individual) and their consequences, and to better understand the penetrance of *P R K N* pathogenic variants, both SmVs and SVs. For Instance, deletion of exons 3-4 was frequently reported as homozygous pathogenic variant in AR *PRKN*-PD patients in MDSGene. However, some studies have suggested that deletions of both exons 3 and 4 maintain the reading frame and are associated with a later onset, milder disease, in contrast to deletion of exon 3 that leads to a frameshift with more severe consequences^28,29^.

Several limitations stem from the current state of knowledge regarding the genetic etiology of the disease. The understanding of pathogenic variants in the *PRKN* gene may be less comprehensive in certain populations that are under-represented in studies, potentially leading to an underestimation of genetic prevalence in those populations. It is important to note that our analysis was limited to reported *PRKN* pathogenic variants observed in early-onset recessive forms of PD patients. Variants predicted to be pathogenic using *in silico* tools and not (yet) reported in patients were not included in our analysis. While adding these variants would further increase the eGP estimates, caution is warranted as it may result in inflated estimates due to the inclusion of false positive pathogenic variants as discussed above.

Other limitations relate to the current knowledge about genetic variation in the general human population and the different ancestries. While massive efforts, like gnomAD, have drastically improved our understanding of the spectrum of genetic variants on the human genome and their frequencies is various ancestries, and have remarkably benefited genetic diagnosis and other aspects of human health, ongoing and future efforts to further improve the cataloguing of human genetic variation in diverse populations will be required to further improve the accuracy of genetic prevalence estimations. Human reference population databases contain genotypic data generated using different sequencing platforms (whole exome sequencing vs whole genomes) and the number of samples from each ancestry are different. This may result in variability in the accuracy of carrier frequency in different populations and, as a consequence, result in variability in the accuracy of eGP between populations. In our analysis, we calculated a per-variant and per-population carrier frequency confidence interval based on the number of individuals sequenced to account for this limitation.

In addition, limited information about SVs is currently available. This is particularly relevant to *PRKN*-PD in which SVs contribute significantly to the disease etiology. While we carefully considered available data sources that capture information about SVs, and incorporated SVs information from gnomAD v2.1 and from jMorp into our analysis, future increases in sample sizes in diverse populations and improvements in technologies to call SVs and assess their consequences on the protein’s function will be required.

It is crucial to also emphasize that the methodology employed in this study was specifically tailored to estimate the genetic prevalence of autosomal recessive *PRKN*-PD caused by biallelic variants in *PRKN*. Complicating the analysis is the fact that *PRKN* is one of the largest gene in the genome, rendering it more prone to genetic variation. Future studies will be imperative to enhance our understanding of the functional consequences of variants in *PRKN* and their role in AR PRKN-PD.

In summary, the methodology used in this study has proven effective in increasing our understanding in the prevalence of multiple rare recessive diseases. As demonstrated through our investigation into *PRKN*-PD, it serves as a powerful tool to complement epidemiology research, offering valuable insights for patient identification strategies and informing the design of clinical trials.

## Data Availability

The study used ONLY openly available human data. All data produced in the present work are contained in the manuscript.

## Acknowledgements

NA

## Data availability

All data generated or analyzed during this study are included in this published article and its supplementary information files.

## Author contributions

DD and EHMW: study design, execution, analysis, writing, and editing of the final version of the manuscript.

CD, and WQ: analysis and review of the final version of the manuscript SC, TH, and JP: review and editing of the final version of the manuscript.

DP, JL and GP: study design, review and editing of the final version of the manuscript.

## Funding

Takeda Development Center Americas, Inc. provided funding.

## Competing interests

DD, CD, JL, SC, TH, JP, DP and GP are employees of Takeda Development Center Americas, Inc.; TH and WQ are employees of Takeda Pharmaceuticals Company Limited. At time of the study, EHMW was an employee of Takeda Development Center Americas, Inc. DD, EHMW, CD, JL, SC, TH, JP, DP and GP are stockholders of Takeda Pharmaceuticals Company Limited. All authors declare no non-financial competing interests.

## Notes

### Author Declarations

The study used ONLY openly available human data. All data sources are listed in the Methods section and include MDSGene (www.mdsgene.org), ClinVar (https://www.ncbi.nlm.nih.gov/clinvar/), gnomAD (https://gnomad.broadinstitute.org) and JMorp (https://jmorp.megabank.tohoku.ac.jp/).

### Summary of Updates

Results section updated to clarify external data used to identify known pathogenic mutations. Revisions do not affect actual results and conclusions.

